# SARS-CoV-2 sample-to-answer nucleic acid testing in a tertiary care emergency department: evaluation and utility

**DOI:** 10.1101/2020.07.03.20145383

**Authors:** Pia Jokela, Anu E Jääskeläinen, Hanna Jarva, Tanja Holma, Maarit J Ahava, Laura Mannonen, Maija Lappalainen, Satu Kurkela, Raisa Loginov

**Author notes:** **Address correspondence to:** Anu E Jääskeläinen, Helsinki University Hospital, HUSLAB, Virology and Immunology, P.O.B. 720 (Topeliuksenkatu 32), FIN-00029 HUS, Finland. Pia Jokela and Anu E Jääskeläinen contributed equally to this work. Author order was determined randomly.

## Abstract

Rapid sample-to-answer tests for detection of SARS-CoV-2 are emerging and data on their relative performance is urgently needed. We evaluated the analytical performance of two rapid nucleic acid tests, Cepheid Xpert^®^ Xpress SARS-CoV-2 and Mobidiag Novodiag^®^ Covid-19, in comparison to a combination reference of three large-scale PCR tests. Moreover, utility of the Novodiag^®^ test in tertiary care emergency departments was assessed. In the preliminary evaluation, analysis of 90 respiratory samples resulted in 100% specificity and sensitivity for Xpert^®^, whereas analysis of 107 samples resulted in 93.4% sensitivity and 100% specificity for Novodiag^®^. Rapid SARS-CoV-2 testing with Novodiag^®^ was made available for four tertiary care emergency departments in Helsinki, Finland between 18 and 31 May, coinciding with a rapidly declining epidemic phase. Altogether 361 respiratory specimens, together with relevant clinical data, were analyzed with Novodiag^®^ and reference tests: 355/361 of the specimens were negative with both methods, and 1/361 was positive in Novodiag^®^ and negative by the reference method. Of the 5 remaining specimens, two were negative with Novodiag^®^, but positive with the reference method with late Ct values. On average, a test result using Novodiag^®^ was available nearly 8 hours earlier than that obtained with the large-scale PCR tests. While the performance of novel sample-to-answer PCR tests need to be carefully evaluated, they may provide timely and reliable results in detection of SARS-CoV-2 and thus facilitate patient management including effective cohorting.

## Introduction

Patients with COVID-19 disease can present with a number of different, and often unspecific signs and symptoms. Thus, the diagnosis of COVID-19 relies on molecular testing of SARS-CoV-2, typically from respiratory specimens (1). Several laboratory-developed and commercial methods are available for this purpose (2-6), both large-scale testing platforms for handling batches of several samples, and simple cartridge-based tests for rapid examination of one or few samples at a time.

Rapid and reliable laboratory testing is essential for patient management and infection control of COVID-19 in closed settings such as hospitals, and it is a prerequisite for appropriate cohorting of patients. Rapid SARS-CoV-2 molecular testing that can be performed near the healthcare facility is urgently needed. A number of such tests have now become available, and variable performance values have been reported for them (7-14).

We aimed to evaluate the analytical performance of two sample-to-answer rapid PCR tests for the detection of SARS-CoV-2 infection, Cepheid Xpert^®^ Xpress SARS-CoV-2 and Mobidiag Novodiag^®^ Covid-19, and to assess the usefulness of such tests at tertiary care emergency departments. Patients who become hospitalized through emergency departments are among those who will benefit the most from test results that are quickly available. Here we describe the utility of a rapid test compared to large-scale testing platforms in such a patient care setting.

## Materials and methods

The study was conducted at the Helsinki University Hospital Laboratory (HUSLAB), Helsinki, Finland. Data were collected and samples treated according to permit HUS/157/2020 (Helsinki University Hospital, Finland).

### Test methods

The evaluated sample-to-answer tests were Cepheid Xpert^®^ Xpress SARS-CoV-2, software version 1.0, later referred to as Xpert^®^, and Mobidiag Novodiag^®^ Covid-19, software version v1.0.1, later Novodiag^®^. Both of these tests are cartridge-based platforms that perform sample preparation, nucleic acid extraction and amplification, as well as detection of the target sequences. The main features of tests are listed in Table 1.

**Table 1.** Features of the evaluated and reference tests. LDT, laboratory developed test; LoD, limit of detection, TCID50, 50 percent tissue culture infective dose.

|  | **Evaluated tests** |  | **Reference tests** |  |  |
| --- | --- | --- | --- | --- | --- |
|  | **Cepheid Xpert® Xpress SARS-CoV-2** | **Mobidiag Novodiag® Covid-19** | **Roche Diagnostics Cobas® SARS-CoV-2** | **Mobidiag Amplidiag® Covid-19 kit** | **LDT** |
| **Intended use** | Nasopharyngeal, oropharyngeal, nasal, or mid-turbinate swab and/or nasal wash/ aspirate | Nasopharyngeal sample | Nasal, nasopharyngeal or oropharyngeal swab samples | Nasopharyngeal sample | Nasal, nasopharyngeal, oropharyngeal swab, nasopharyngeal/tracheal aspirate, sputum or faeces. |
| **Sample volume µl** | 300 | 250 | 350 | 360 | 250 |
| **Targets** | E and N2 | orf1ab and N | orf1ab and E | orf1ab and N | N |
| **Internal controls** | Sample processing control, probe check control | Sampling control, process control | Process control | Sampling control | no |
| **External controls** | Not included, but recommended by the manufacturer | Not included, but recommended by the manufacturer | Negative control and a low titer positive control | Negative and positive controls | 2 negative controls (H<sub>2</sub>O, no-template control), positive control |
| **LoD, as reported by the manufacturer** | 250 c/ml | 313 c/ml | 0,009 TCID<sub>50</sub>/ml (ORF1/a)/ 0,003 TCID<sub>50</sub>/ml (E) | 1250 c/ml |  |
| **Assay run time** | ~45 min | ~1h 15 min | ~3 h | ~2,75h | ~2h + extraction ~1h |

The three platforms used in our laboratory for routine diagnostics of SARS-CoV-2 were deployed as reference tests: the WHO recommended laboratory-developed test (LDT), modified from Corman and others (2) [Corman et al., 2020], cobas^®^ SARS-CoV-2 test kit on the cobas^®^ 6800 platform (Roche Diagnostics, Basel, Switzerland), and Amplidiag^®^ COVID-19 test on the Amplidiag^®^ Easy platform (Mobidiag, Espoo, Finland). We have separately evaluated the performance of the three reference methods used in our laboratory, and shown a good agreement between them (Mannonen and others, unpublished). The clinical performance of the latter two has been established by the manufacturers. See Table 1 for the main features of the tests.

### Patient samples and proficiency samples for the analytical evaluation

Altogether 107 nasopharyngeal or oropharyngeal swab specimens sent to HUSLAB were included in the evaluation: all of them were tested with Novodiag^®^, and 90 with Xpert^®^. Of the 107 specimens, 97 were sent for SARS-CoV-2 testing between March and May 2020, and 10 were sent due to suspicion of other respiratory virus infection in 2019 or early 2020.

Sixty-one were SARS-CoV-2 positive and 46 negative in the reference SARS-CoV-2 PCR tests. All specimens were analyzed by at least one of our reference tests. Those specimens that gave discrepant results were analyzed with at least the cobas^®^ SARS-CoV-2 test.

Of the 10 samples originally sent for other than SARS-CoV-2 testing, 8 were originally tested by Allplex Respiratory Panel 1/2/3 (Seegene, Seoul, Republic of Korea) and two by xTAG RVP Fast (Luminex Diagnostics, Toronto, Canada). One was positive for human coronavirus (CoV) OC43, one for CoV 229E and human rhinovirus. The remaining samples were positive for other potentially interfering respiratory viruses: parainfluenzavirus 1 (1 sample), parainfluenza virus 2 (1), parainfluenza virus 3 (1), adenovirus (1), human metapneumovirus (1), human rhinovirus and bocavirus (1), respiratory syncytial virus (RSV; 1), influenza virus A (pdm09; 1) and influenza virus B (1).

In addition, altogether 8 proficiency samples of the QCMD 2020 Coronavirus Outbreak Preparedness EQA Pilot Study (Glasgow, Scotland, UK) containing CoVs SARS-CoV-2 (5 samples), OC43 (1) and NL63 (1) were analyzed to evaluate the performance of the Novodiag^®^ test.

### Patient samples from emergency departments

This study was conducted with specimens from the emergency departments of the following tertiary care hospitals in Helsinki, Finland: Meilahti Tower Hospital, Haartman Hospital, Malmi Hospital, and the New Children’s Hospital. Altogether, 362 nasopharyngeal specimens were sent to our laboratory for rapid PCR testing from these emergency departments between 18 to 31 May 2020, which coincided with a declining epidemic phase of COVID-19 in Finland. Three of these emergency departments are located within 1 km from our laboratory, where the testing was performed, and one is located at a distance of 13 km. It was agreed that rapid testing would be primarily targeted for those patients who were likely to be hospitalized, and tests would be requested according to clinical assessment. The specimens first underwent rapid PCR testing with Novodiag^®^, and the result was immediately reported. Directly after pipetting the Novodiag^®^ cassette, the specimen was subjected to one of the three reference tests for confirmation and for the purposes of clinical evaluation.

### Statistical analysis

Concordance of the results obtained by the Novodiag^®^ and Xpert^®^ assays in comparison to a combination reference of the three large-scale PCR tests was examined in McNemar’s test. Statistical significance was set at P < 0.05. To assess the agreement between the methods by chance, Cohen’s kappa coefficient (κ) was computed. Mann-Whitney U test was used to compare the Ct value medians, and the turnaround times of Novodiag^®^ and the three large-scale PCR tests was examined using Wilcoxon signed ranks test. Statistical analysis was performed using SPSS/PASW statistical program package, version 25 (IBM SPSS Inc., Chicago, IL, USA).

## Results

### Analytical evaluation

The results of the analytical evaluation are summarized in Table 2. The performance of the Xpert^®^ and Novodiag^®^ tests was assessed in analysis of 90 and 107 upper respiratory samples, respectively, in comparison to one of the three reference PCR tests. Respiratory specimens with a positive result in one of the reference methods were considered as true positives. Four of the samples were analyzed with two reference tests. The Xpert^®^ assay yielded a valid result for all specimens, and they were 100% (90/90 specimens) consistent compared to the reference with kappa value of 1.000 (P < 0.001). The Novodiag^®^ assay yielded a valid result for all but one specimen (invalid rate of 0.93%) and an agreement of 96.2 % (102/106), kappa value of 0.924 (P < 0.001) with the reference. No difference of statistical significance between the results of the Novodiag^®^ and the reference tests was found (P=0.125). For the samples positive with cobas^®^ SARS-CoV-2 test, the median Ct value of SARS-CoV-2 spesific target 1 was significantly higher in the four samples with discordant results (31.9) than that for the 19 concordant results (21.6, P=0.002). Discrepant samples are listed in Table 3. In analysis of the proficiency samples, Novodiag^®^ failed to detect two SARS-CoV2 containing samples with 3.3 log_10_ and 2.3 log_10_ copies/ml, the lowest concentrations in the panel.

**Table 2a.**
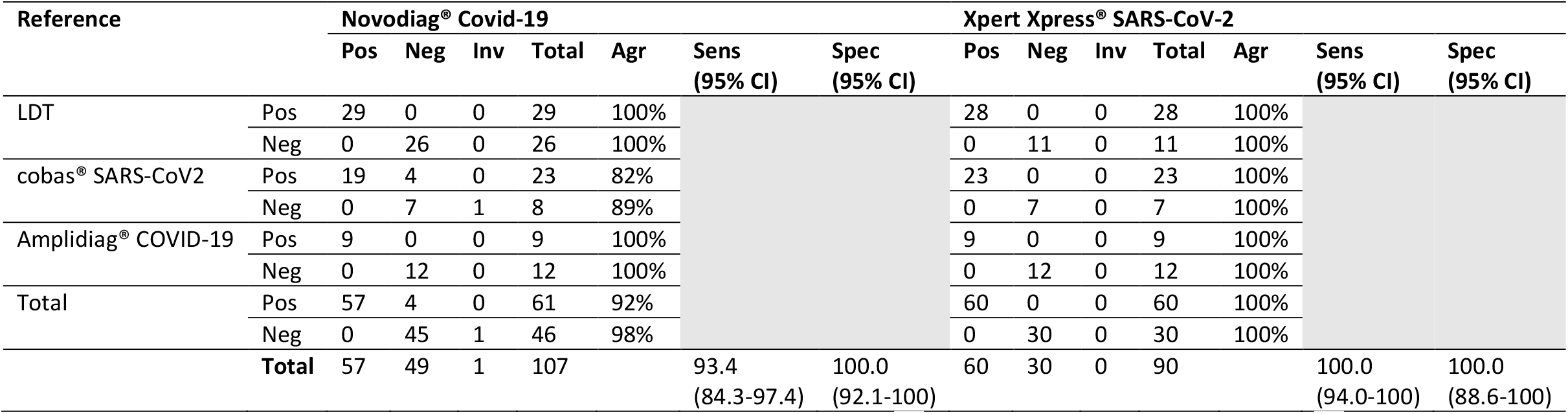
Number of tested samples and performance of the Novodiag® Covid-19 assay and the Xpress® SARS-CoV-2 assay in the initial evaluation. LDT; laboratory developed test; Pos, positive; Neg, negative, Inv, invalid, CI, confidence interval.

**Table 2b.**
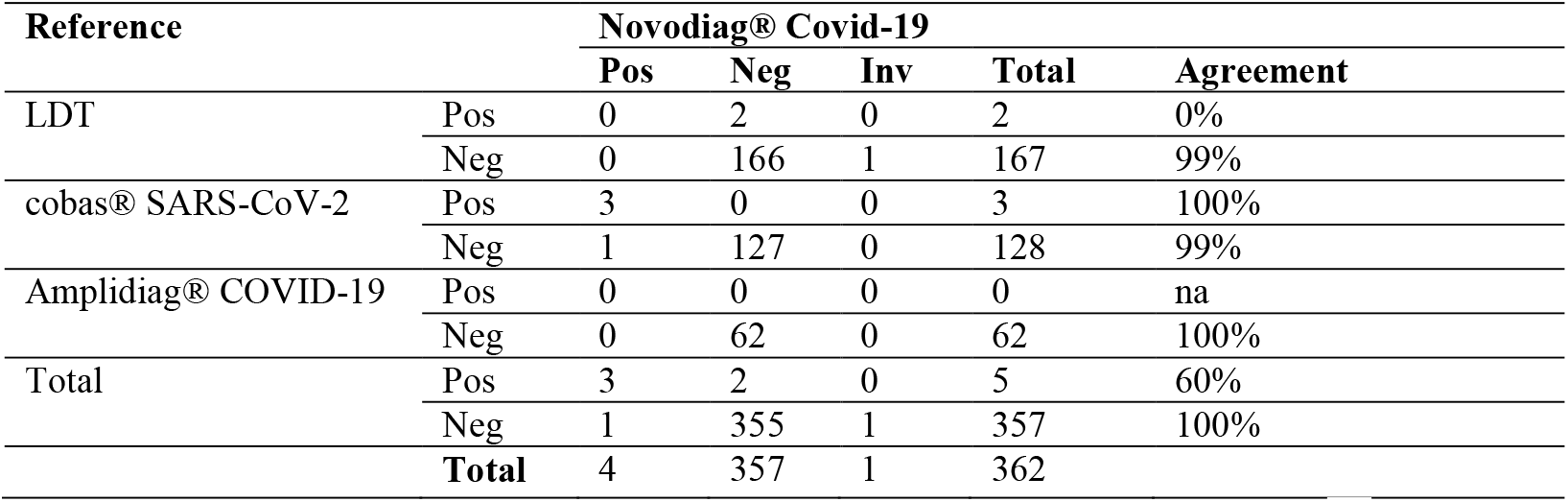
Number of tested samples and performance of the Novodiag® Covid-19 assay at the tertiary care emergency department evaluation. LDT; laboratory developed test; Pos, positive; Neg, negative, Inv, invalid; na, not applicable.

**Table 3.** Discrepant samples. Samples 1-5 on white background: samples that gave discrepant result using Novodiag^®^ and reference test in the initial evaluation. Samples 6-9 on gray background: samples that gave discrepant result using Novodiag^®^ and reference test in the emergence department utility evaluation. Sample 3 was reference tested by both cobas and Amplidiag tests. Sample 5 was reference tested by both cobas and LDT. ^1^Reference test Ct values when sample positive in reference test: for cobas^®^: target 1/ Target 2, for Amplidiag^®^: N/orf1ab, for LDT: N. LDT, laboratory developed test, ND, not detected.

| Reference |  |  |  | Novodiag<sup>®</sup> COVID-19 |  |  | Xpert<sup>®</sup> Xpress SARS-CoV-2 |  |  |
| --- | --- | --- | --- | --- | --- | --- | --- | --- | --- |
| Sample | Test | Result | Ct<sup>1</sup> (pos samples) | Result | N | orf1ab | Result | Ct E (pos samples) | Ct N (pos samples) |
| 1 | cobas<sup>®</sup> SARS-CoV-2 | Posit | 33.15/36.77 | Negat | Negat | Negat | Posit | 0 | 39.9 |
| 2 | cobas<sup>®</sup> SARS-CoV-2 | Posit | 32.32/34.46 | Negat | Negat | Negat | Posit | 0 | 39.8 |
| 3 | cobas<sup>®</sup> SARS-CoV-2 | Posit | 31.53/33.41 | Negat | Negat | Negat | Posit | 32.1 | 35.6 |
|  | Amplidiag<sup>®</sup> COVID-19 | Posit | 38.1/0 |  |  |  |  |  |  |
| 4 | cobas<sup>®</sup> SARS-CoV-2 | Posit | 28.14/28.09 | Negat | Negat | Negat | Posit | 27.3 | 30 |
| 5 | cobas<sup>®</sup> SARS-CoV-2 | Negat |  | Invalid |  |  | ND |  |  |
|  | LDT | Negat |  |  |  |  |  |  |  |
| 6 | LDT | Posit | 37.62 | Negat | Negat | Negat | Negat |  |  |
| 7 | LDT | Posit | 38.48 | Negat | Negat | Negat | Negat |  |  |
| 8 | cobas<sup>®</sup> SARS-CoV-2 | Negat |  | Posit | Negat | Posit | Negat |  |  |
| 9 | LDT | Negat |  | Invalid |  |  | ND |  |  |

Other respiratory viruses, including seasonal coronaviruses OC43 and 229E, did not cause any false positive test results in this material including patient and proficiency samples.

### Evaluation of the utility of the Novodiag^®^test with specimens from tertiary care emergency departments

The clinical evaluation included 362 samples analyzed by Novodiag^®^ and reference PCR test from 356 patients attending tertiary care emergency departments. One specimen was excluded from further analyses due to invalid sampling control detected by Novodiag^®^. The remaining 361 samples were collected from 356 patients with a median age of 72 years. The patients consisted of 34 children (median age of 5 years) and 322 adults (median age of 74 years), with ages ranging from 2 weeks to 16 years and 19 to 99 years, respectively.

Altogether, 356/361 samples were negative according to a reference test. One of these was positive with Novodiag^®^, so the specificity of Novodiag^®^ in this setting was 99.7%. Of the five reference-test positive samples, three were positive by Novodiag^®^. The two false-negatives by Novodiag^®^ had high Ct values of N gene target in the reference LDT (37.62 and 38.48). The discrepant samples are listed in Table 3.

The prevalence of SARS-CoV-2 PCR positivity among these patients, including people attending tertiary care emergency departments due to other reasons than suspicion of COVID-19, was 1.4%. Fever, respiratory and gastrointestinal symptoms were recorded at attendance and are summarized in Table 4. 86 patients had no clear symptoms pointing towards COVID-19. Patients positive for SARS-CoV-2 according to a reference test were

**Table 4.**
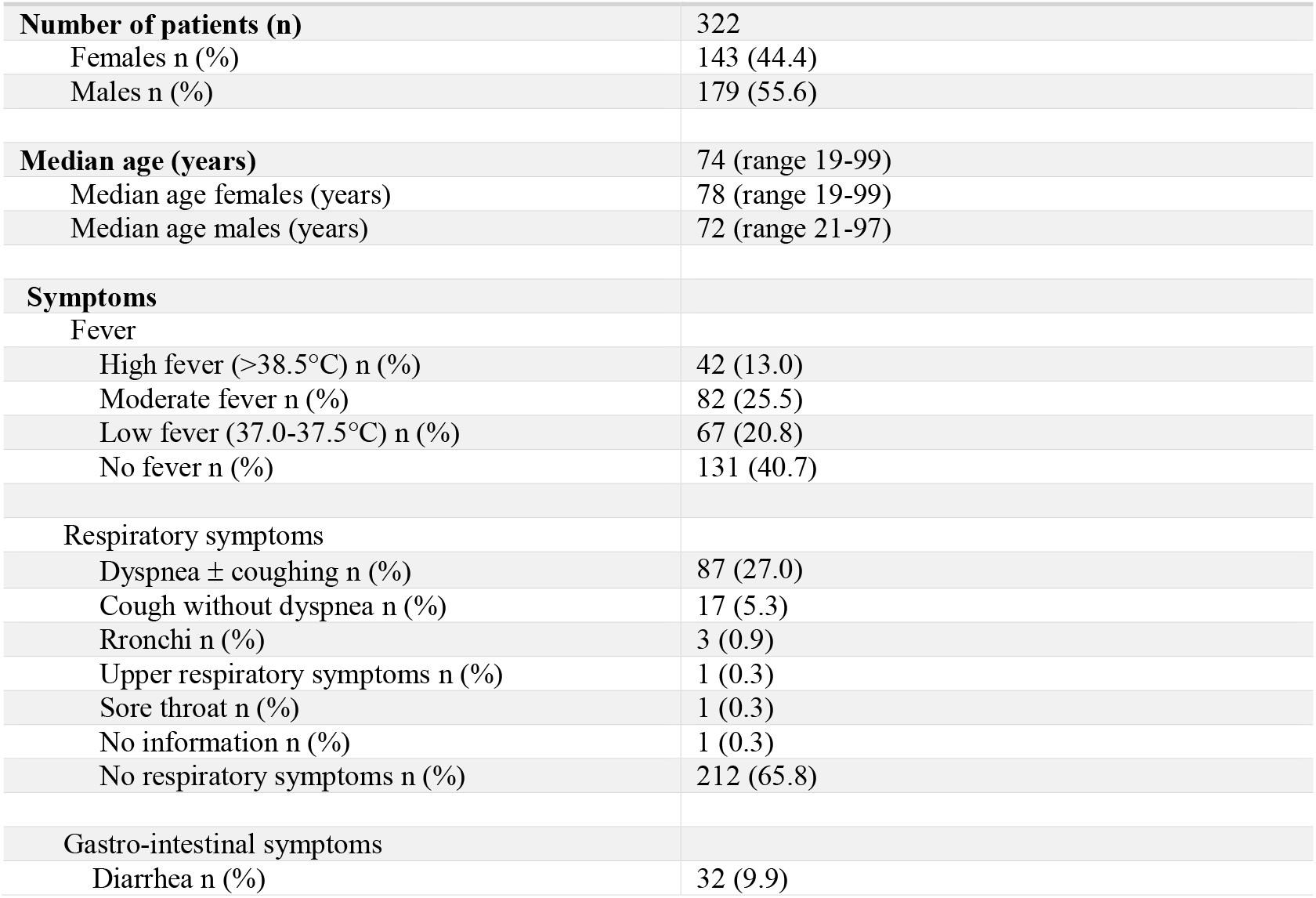
Clinical characteristics of the adult patients at the first evaluation at the emergency department. Some patients presented with multiple symptoms.

|  |  |
| --- | --- |
| **Number of patients (n)** | 322 |
| Females n (%) | 143 (44.4) |
| Males n (%) | 179 (55.6) |
| **Median age (years)** | 74 (range 19-99) |
| Median age females (years) | 78 (range 19-99) |
| Median age males (years) | 72 (range 21-97) |
| **Symptoms** |  |
| Fever |  |
| High fever (>38.5°C) n (%) | 42 (13.0) |
| Moderate fever n (%) | 82 (25.5) |
| Low fever (37.0-37.5°C) n (%) | 67 (20.8) |
| No fever n (%) | 131 (40.7) |
| Respiratory symptoms |  |
| Dyspnea ± coughing n (%) | 87 (27.0) |
| Cough without dyspnea n (%) | 17 (5.3) |
| Rronchi n (%) | 3 (0.9) |
| Upper respiratory symptoms n (%) | 1 (0.3) |
| Sore throat n (%) | 1 (0.3) |
| No information n (%) | 1 (0.3) |
| No respiratory symptoms n (%) | 212 (65.8) |
| Gastro-intestinal symptoms |  |
| Diarrhea n (%) | 32 (9.9) |

1. 97-year-old male, slight temperature (37°C), reduced general condition. The patient had no respiratory or gastro-intestinal symptoms.
2. 45-year-old male, high fever (39.9°C) and shortness of breath.
3. 84-year-old male, no fever, no respiratory of gastro-intestinal symptoms. The patient had fallen at home and was brought to hospital. The patient died after a week in the hospital. No intensive care was begun because of the patient’s Alzheimer’s disease.
4. 71-year-old male, slight temperature (37°C), shortness of breath and coughing and diarrhea. This patient had several underlying diseases (type II diabetes, liver cirrhosis, hypertension, psoriasis) and he succumbed to the infection after 2 days in the hospital. Intensive care was not considered useful.
5. 16-year-old male, whose medical data is not available.

The one patient who was positive for SARS-CoV-2 according to Novodiag^®^ but negative in a reference test, was a 53-year-old man with acute myocardial infarction, no fever or other symptoms suggesting COVID-19.

All of the clinical evaluation samples were analyzed with one of the reference tests on large-scale PCR platforms immediately after the Novodiag^®^ cassette was pipetted. The Novodiag^®^ assay resulted in statistically significant acceleration of diagnostics by enabling results with median turnaround time of 3 h 54 min as compared to the median turnaround time of 11 h 44 min obtained with the reference tests (P<0.001). These times include transportation of the samples from the emergency departments to the laboratory.

## Discussion

The current epidemiological situation underlines the need for rapid SARS-CoV-2 diagnostic tests with comparable performance with the more time-consuming, technically demanding and labor-intensive tests currently employed by many laboratories. Implementing rapid, easy to use diagnostic approaches is especially important in remote locations where distance to specialized microbiological laboratories may cause severe delays in specimen transportation and diagnosis of COVID-19 patients. The evaluated cartridge-based rapid PCR tests, Xpert^®^ Xpress SARS-CoV-2 and Novodiag^®^ Covid-19, provide automated analysis of results and storage of data therefore reducing the level of expertise and information system management required. Moreover, the tests are on random-access platforms and require no batching of samples or processing of sample prior to analysis, therefore offering relatively short turnaround times with minimal equipment.

In the analytical performance assessment of this study, Xpert^®^ showed complete concordance of results with the reference and the kappa value of 1.00 implied an almost perfect agreement. The high sensitivity and specificity of 100% observed for Xpert^®^ in this study has also been shown in previous reports (7-9).

For Novodiag^®^, the results obtained were 96.2% concordant with the reference. The kappa value of 0.924 also referred to an almost perfect agreement, which was further supported by McNemar’s test. Together with the two false negative low-concentration proficiency samples, the high median Ct value of 31.9 for the false negative patient samples may point towards a constricted ability of the Novodiag^®^ to detect positive samples with low viral loads (Tables 3 and 5). Nonetheless, with high specificity of 100% and sensitivity of 93.4%, and low invalid rate of 0.9%, the Novodiag^®^ Covid-19 test was chosen for the utility assessment of rapid SARS-CoV-2 testing in the clinical setting of emergency departments. Due to the inactivation protocol included in the sample preparation step of the Novodiag^®^ Covid-19 test, there is no need for placing the Novodiag^®^ instrument inside a biosafety cabinet. This, together with potentially foreseeable challenges in the availability of Xpert^®^ test cassettes, encouraged us to choose Novodiag^®^ for the clinical utility study.

**Table 5.** Performance of the Novodiag® Covid-19 assay and the Xpress® SARS-CoV-2 assay in the preliminary evaluation.

| Reference | Novodiag® Covid-19, n=106 |  | Xpress® SARS-CoV-2, n=90 |  |
| --- | --- | --- | --- | --- |
|  | No/reference | Agreement % (95% CI) | No/reference | Agreement % (95% CI) |
| Posit | 57/61 | 93.4 (84.3-97.4) | 60/60 | 100 (94.0-100) |
| Ct <20 | 15/15 | 100.0 (79.6-100) | 15/15 | 100.0 (79.6-100) |
| Ct 20-30 | 38/39 | 97.4.0 (86.8-99.5) | 38/38 | 100.0 (90.8-100) |
| Ct >30 | 4/7 | 57.1 (25.0-84.2) | 7/7 | 100.0 (64.6-100) |
| Negat | 45/45 | 100.0 (92.1-100) | 30/30 | 100.0 (88.6-100) |

The utility of rapid SARS-CoV-2 testing with Novodiag^®^ was assessed prospectively in the analysis of 362 samples from four tertiary care emergency departments in Helsinki, Finland, between 18 and 31 May, 2020. At that time, the number of new cases was quickly declining (on the average 28 cases per day in the Helsinki and Uusimaa hospital district (incidence 11.6 /100 000)) (15) and approximately 2% of all specimens sent to HUSLAB were positive (unpublished data). As expected, a slightly lower positivity rate of 1.4 % was observed for the patients at the emergency departments, the majority of whom sought health care services primarily due to reasons other than those suggesting COVID-19. Together with the clinical profiles of the positive patients, this epidemiological snapshot on the frequency of COVID-19 infections showed cases of COVID-19 patients with unspecific clinical picture. This emphasizes the need for a rapid SARS-CoV2 testing in emergency departments and hospital settings.

As a response to the current need for extensive SARS-CoV-2 testing, several commercial nucleic acid detections assays have become available, all of which have their advantages and limitations. In the present study, the Novodiag^®^ Covid-19 with reasonable sensitivity and high specificity in the analytical evaluation was made available for rapid SARS-CoV2 testing for four emergency departments. Timely results are required to facilitate efficient patient flow and effective patient cohorting. Subclinical COVID-19 infections do occur (16) and could potentially lead to spread of the disease in hospital settings. Prompt diagnosis is of especially high priority for patients admitted to the hospital through the emergency departments and therefore the possibility for rapid SARS-CoV-2 testing was first offered to these settings. Indeed, implementation of the Novodiag^®^ provided results nearly 8 hours faster as compared to the large-scale PCR tests.

A limitation of the present study is the use of three different large-scale PCR tests for reference, as analysis of all samples on one platform was not possible due to the global shortage in testing supplies at the time of the study and a heavy load of samples to be tested. As a limitation, it should also be noted that age distribution and clinical information was available only for the samples of the clinical evaluation.

In conclusion, the Xpert^®^ Xpress SARS-CoV-2 showed high sensitivity and specificity, and a reasonable sensitivity and high specificity was achieved for the Novodiag^®^ Covid-19 assay. The possible limited ability of the Novodiag^®^ Covid-19 test to detect low viral load samples is a drawback, which may be overcome by confirmatory testing depending on the clinical context. Taken together, with the acceleration of diagnostics and the ease of use, rapid sample-to-answer PCR tests may provide timely and reliable results with a positive impact on the management of patient flow and infection control in the prevention of nosocomial COVID-19 infections.

## Data Availability

Data is either included in the manuscript or if missing, available upon request.

## Acknowledgements

We would like to thank the nursing staff at our laboratory, especially Jonna Keijama, for the valuable technical assistance.

## Author contributions

Conceptualization: PJ, AEJ, SK, ML, HJ, RL. Data curation: PJ, AEJ, MA, HJ, RL. Formal Analysis: PJ, AEJ, HJ, RL. Funding acquisition: ML. Investigation: PJ, AEJ, MA, HJ, RL. Methodology: PJ, AEJ, TH, LM, RL. Project administration: PJ, AEJ, ML, RL. Resources: ML. Supervision: RL. Validation: PJ, AEJ, TH, RL. Visualization: PJ, AEJ, HJ, RL. Writing – original draft: PJ, AEJ, SK, HJ, RL. Writing – review & editing: PJ, AEJ, TH, MA, SK, LM, ML, HJ, RL

